# Choice of Epilepsy Anti-Seizure Medications and Associated Outcomes in Medicare Beneficiaries

**DOI:** 10.1101/2025.03.18.25324227

**Authors:** Julianne D. Brooks, Rafaella Cazé de Medeiros, Shuo Sun, Madhav Sankaranarayanan, M. Brandon Westover, Lee H. Schwamm, Joseph P. Newhouse, Sebastien Haneuse, Lidia M.V.R. Moura

## Abstract

**Background:** The lack of specific guidelines for seizure treatment after acute ischemic stroke (AIS), makes the choice of an appropriate anti-seizure medication choice a challenge for providers because each drug may have different adverse effects and outcomes.

**Methods:** In this retrospective matched cohort study, we analyzed a 20% sample of U.S. Medicare beneficiaries aged 65 and over hospitalized for a first acute ischemic stroke (AIS) between 2009-2021 who were discharged home. We included individuals who were enrolled in Medicare hospital, medical and prescription drug insurance for 12 months prior to hospitalization and were not taking epilepsy-specific anti-seizure medication (ESM) prior to hospitalization. We matched individuals on days from discharge to ESM initiation. Individuals who initiated ESMs other than Levetiracetam, i.e. Lamotrigine, Carbamazepine, Oxcarbazepine within 30 days of discharge (N = 229) were matched to Levetiracetam initiators (N =687). We investigated the time to seizure-like events, emergency department (ED) visits, and re-hospitalizations with a follow-up of 180 days after initiation using a semi-competing risk framework. We estimated the average treatment effect among the treated i.e. those who received other ESMs.

**Results:** The matched cohort of 916 ESM initiators had a median age of 74 (IQR 69, 82) and was 57% female and 71% Non-Hispanic White. Using the semi-competing risk framework, those who received other ESM had a 37% lower hazard of seizure-like events compared to receiving LEV, given that death had not occurred, hazard ratio 0.63 (95% CI: 0.43, 0.91). Among those who initiated ESMs other than Levetiracetam, the hazard of ED visits and hospitalizations, given that death had not occurred, did not different significantly from initiating Levetiracetam; hazard ratios 1.00 (95% CI: 0.80, 1.25) and 0.98 (95% CI: 0.75, 1.28), respectively.

**Conclusion:** In a sample of Medicare beneficiaries hospitalized for acute ischemic stroke and discharged home, initiating Levetiracetam in the outpatient setting was associated with a higher risk of seizure-like events compared to other ESMs. However, no significant differences were observed in the incidence of ED visits or hospitalizations, suggesting comparable safety profiles in these broader clinical outcomes.

## INTRODUCTION

Stroke is the most common cause of seizures in older adults.^1^ Epilepsy-specific anti-seizure medications (ESMs) are used for post-stroke seizure management.^2,3^ Several ESM options are available, including Levetiracetam (LEV), Lamotrigine (LTG), Valproate (VPA), Lacosamide (LAC), and others. Studies indicate that second-generation ESMs, such as LEV and LTG, and third-generation options, including LAC, are better tolerated in adults 65 and over compared to first-generation drugs like phenytoin (PHT) and carbamazepine (CBZ).^4,5^ These newer classes of ESMs are associated with fewer drug-drug interactions and adverse outcomes, making them more suitable for older populations.

However, despite these advancements, there remains a critical gap in understanding the comparative effectiveness and safety of second- and third-generation ESMs in older adults recovering from stroke. Current evidence often focuses on younger populations, leaving older patients underrepresented.^3,6,7^ This is especially concerning given the well-documented adverse effects of ESMs, including dizziness, fatigue, unsteadiness, and mood or behavioral changes.^7,8^ For older stroke survivors, these side effects can exacerbate existing functional impairments and neuropsychiatric conditions. Bridging this evidence gap is vital to guide clinicians in selecting the most effective and safest ESMs following acute ischemic stroke (AIS) for this vulnerable population, optimizing recovery outcomes and quality of life.^9,10^

Among Medicare beneficiaries prescribed antiseizure medications, LEV is the most commonly initiated drug for new-onset seizures and is often used as monotherapy.^11,12^ Its popularity is largely attributed to its ease of administration (available in both intravenous and tablet forms, with no titration required) and minimal drug-drug interactions.^13,14^ These characteristics are particularly important for elderly stroke survivors, who often have multiple comorbidities and are prescribed anticoagulants, antiarrhythmics, or antihypertensive medications that may interact with other ESMs.^15,16^

While LEV’s profile makes it a practical choice, known side effects such as drowsiness and impaired balance can lead to falls, a critical concern in this population.^2,26^ Falls often result in ED visits, hospitalizations, and loss of independence, highlighting the heightened vulnerability of older ambulatory stroke survivors discharged home - a less supervised yet highly at-risk group.^17^ Despite this, there is limited evidence examining the impact of ESM choice on adverse outcomes such as unsteadiness or seizure-like events in community-dwelling older stroke survivors. To address these gaps, we investigated the relationship between outpatient ESM choice within 30 days of stroke discharge and subsequent healthcare utilization and seizure-like events in AIS survivors aged 65 and older discharged home.

## METHODS

This study was approved by the Mass General Brigham Institutional Review Board and followed the Strengthening the Reporting of Observational Studies in Epidemiology (STROBE) reporting guidelines.^18^ The requirement for informed consent was waived in our study as we performed a secondary analysis of data routinely collected for billing. The data supporting this study’s findings were collected by The Centers for Medicare & Medicaid Services (CMS) and were made available by CMS with no direct identifiers.^19^ All results were aggregated following CMS Cell Suppression Policies. Restrictions apply to the availability of these data, which were used under license for this study. Medicare data are available through CMS with their permission. We included the code which produced the findings in the supplemental materials (Supplementary materials – analytical code).

### Study Design

We conducted a retrospective analysis of U.S. administrative claims data using a matched cohort study design.^20^ Our focus was on individuals over 65 years old, since this population is more vulnerable to adverse outcomes because of its greater frequency of multiple comorbidities.^21^ We analyzed a 20% sample of U.S. Medicare health insurance beneficiaries, including only adults aged 65 and over discharged home after a hospitalization for AIS between January 1, 2009, and September 30, 2021. Hospitalizations were selected from the Medicare Provider Analysis and Review (MedPAR) database based on principal diagnosis codes for AIS. We selected International Classification of Diseases, 9th Revision (ICD-9) codes 433, 434, 436, and ICD-10 codes I63, a validated strategy to capture AIS in administrative databases.^22^

We included Medicare beneficiaries who were enrolled in the traditional Medicare Part A (hospital insurance), Part B (medical insurance), and Part D (drug prescription coverage) continuously for 12 months before their admission for stroke. We included first stroke admission, using at least 1 year of look back to identify prior stroke.^23^ We focused on outpatient medication initiation and outcomes in community-dwelling stroke survivors. We included individuals who were discharged home and did not include those who were discharged to a skilled nursing facility or other inpatient facility.

We excluded Medicare beneficiaries with an ESM prescription in the 120-day period before hospitalization, to remove prevalent ESM users. A 120-day look-back period accounts for patients with prescriptions of 90-day supplies, as well as a 30-day grace period in order to capture stockpiling of earlier medication refills. We focused on ESM monotherapy, so patients prescribed more than one ESM were excluded. Additional details on sample selection are included in Supplemental Material – Supplemental Methods and Table S1.

### Participant Characteristics

We described the following demographic characteristics for the sample: age, sex, race and ethnicity. Reported race and ethnicity (Non-Hispanic White, Non-Hispanic Black, Hispanic, Asian, Other) were categorized using the Research Triangle Institute (RTI) race/ethnicity variable provided in Master Beneficiary Summary File (MBSF).^24^

We identified baseline comorbid conditions using Medicare’s Chronic Condition Warehouse (CCW).^25^ We considered a patient to have a condition at baseline if they met the diagnosis definition prior to their stroke admission. In addition, we reported the presence of a claim with an ICD code for arrhythmia (ICD-9 code 427.x and ICD-10 codes I48.x and I49.x) within the 12 months prior to admission, as certain ESMs are less appropriate for individuals with specific types of arrhythmia (e.g., lacosamide and its dose-dependent association with atrioventricular block and PR interval prolongation).^26^ We identified beneficiaries with baseline dementia using a validated definition of AD/ADRD, which had excellent accuracy as demonstrated by a CV-AUC of 0.94.^27^ The validated definition of dementia was developed using ICD-10 codes, so we used a previously published crosswalk of ICD-10 to ICD-9 codes to identify beneficiaries with dementia with ICD-9 claims (Supplemental Table S2).^28^

Using claims data, we captured clinical factors that may influence ESM choice and are associated with seizure outcomes and healthcare utilization: Stroke severity, which correlates with higher modified Rankin Scores (mRS)^29^, is a well-established risk factor for late seizures.^30^ Additionally, individuals with higher mRS scores at discharge are more likely to have unfavorable outcomes, which may lead to ED visits and hospitalizations. The mRS has seven total categories ranging from no or low disability to death: 0 (no symptoms), 1 (no significant disability), 2 (slight disability), 3 (moderate disability), 4 (moderate to severe disability), 5 (severe disability), 6 (death).^31,32^ For this study, we used a validated claims-based algorithm to classify mRS as a binary outcome, grouping scores of 0 to 3 and 4 to 6. The algorithm was able to accurately identify disability status with an ROC AUC of 0.85.^33^

### Matching Characteristics

A common issue in real-world time-to-event analysis is immortal time bias due to misaligned treatment start.^34^ Our sample was selected on initiation of treatment within 30 days of discharge, but healthcare utilization and outcomes may vary within this window. To address this, we utilized a matched cohort design, matching initiators in our comparison groups on days to initiation.^35^

The ESM initiators were defined as the beneficiaries with therapy initiation within 30 days of post-stroke discharge, beginning at the index acute hospitalization discharge date. Prescription claims were identified in the Medicare Part D prescription data using the generic and commercial brand names listed in Supplemental Table S3. We used an intention-to-treat strategy, so we categorized individuals based on whether they were prescribed a drug, indicated by a prescription claim for ESM.

We grouped ESM initiators into two groups for matching: LEV initiators and other ESM Initiators, which includes LTG, LAC, CBZ, VPA and other ESMs. For each beneficiary in use of other ESM, we identified 3 matches undergoing treatment with LEV. We calculated the Mahalanobis distance from other ESM Initiators to LEV initiators based on days from discharge to medication initiation and selected matches based on the shortest distance.^36^ In this matching process patients on LEV were used as controls for each patient using another ESM (matching without replacement).

### Outcomes

The outcomes measured were the time to seizure-like events, time to hospital readmissions, and ED visits with a follow up period of 180 days after initiation. We treated mortality as a competing risk.

We measured the time to seizure-like events using claims with diagnosis codes for seizure-like events from the inpatient, outpatient and carrier claims files. A list of seizure diagnosis codes was identified from the CCW definition of epilepsy and is included in Supplemental Table S4. We considered individuals to meet the definition for a seizure-like event if they had one inpatient claim or two outpatient claims occurring more than one day apart. We considered the time to the first day the individual met the criteria.

We identified hospital readmissions using acute hospitalizations claims occurring after the index stroke hospitalization in the MedPAR file. ED visits were identified using outpatient and inpatient claims with revenue center code indicative of an ED visits (0450, 0451, 0452, 0456, 0459, 0981).^37^

We used the beneficiaries date of death [BENE_DEATH_DT] from the Medicare MBSF, which comes from several sources including the Social Security Administration. Overall 99% of the death information in the MBSF has been validated.^38^

Individuals were followed from the day of medication initiation to the first occurrence of the outcome, mortality or a censoring event. Censoring events included the end of the study observation period (180 days after initiation). Using Medicare claims data, we were able to follow individuals to the end of the study period.

### Statistical Analysis

Our matching method allows us to estimate the average treatment effect in the treated group, the Other ESM group. In other words, we estimated the effect of receiving treatment with other ESMs compared with those individuals receiving LEV.

We used a semi-competing risks framework to analyze time to seizure like events, ED visits, and hospital readmission with mortality as a competing risk. The semi-competing risks framework is described previously in Haneuse and Lee 2016.^39^ In brief, this framework allows us to consider the occurrence of a non-terminal event (seizure like events, readmission, ED visit), which is subject to a terminal event (death). We used an illness-death model, which defines 3 hazard functions. The first hazard function is for seizure-like events, given that neither seizure-like events nor death has occurred. The second hazard function is for death, given that neither seizure-like events n nor death has occurred. The third function is for time to death after seizure like event has occurred. Baseline hazard functions for all survival models were structured using B-splines. We presented results for the illness-death model based on the semi-Markov specification. We used the SemiCompRisk and SemiCompRiskFreq package for R.

Models were adjusted for days to initiation, age, modified Rankin score, and baseline depression. We did not have missing data for the covariates included in the model. A table with covariates incrementally added to the model specification can be found in the Supplemental Material. We adjusted for age and modified Rankin score as these are related to the outcomes of interest, but did not meet the definition of confounding because they were not related to the exposure. We used a 0.2 threshold for defining a substantial difference based on standardized mean differences.

### Secondary Pre-planned Analysis

We also repeated our analysis, stratifying by age less than or equal to 75 and greater than 75. For this analysis, we did not include medications with fewer than 10 initiators.

### Sensitivity Analysis

A potential concern is the misclassification of seizure-like events when using claims data. To assess the extent to which our results might vary with different definitions of the same construct, we conducted sensitivity analyses using alternative methods to identify seizure-like events. First, we applied the standard CCW definition, which requires one inpatient claim or two outpatient claims suggestive of epilepsy, occurring on different dates. Second, we employed a more sensitive approach that included any inpatient or outpatient claim for seizure-like events irrespective of date. Third, we tested the inclusion of additional ICD codes for seizure-like events, incorporating less specific codes for convulsions. These alternative definitions and their impact on the results are detailed in Supplementary material (Table S5).

## RESULTS

Descriptive statistics for the sample are provided in Table 1. The 229 other ESM initiators were matched to 687 Levetiracetam initiators, for a total sample size of 916 individuals. The study sample was 57% female, had a median age of 74 years old (interquartile range 69 to 81) and was 71% Non-Hispanic White. The percentage of individuals with Alzheimer’s disease and related dementia (AD/ADRD) in the matched sample was 8% for Other ESM and 4% for Levetiracetam initiators (SMD: 0.16). Depression was present at baseline in 60% of Other ESM vs in 36% of Levetiracetam initiators (SMD: 0.5). At discharge, 50% of Other ESM and to 42% of Levetiracetam initiators had an mRS indicative of moderate to severe disability (SMD: 0.18). In the 12 months prior to admission, 34% of LEV and 38% of Other ESM initiators had a claim for arrhythmia (SMD: 0.11).

**Table 1.**
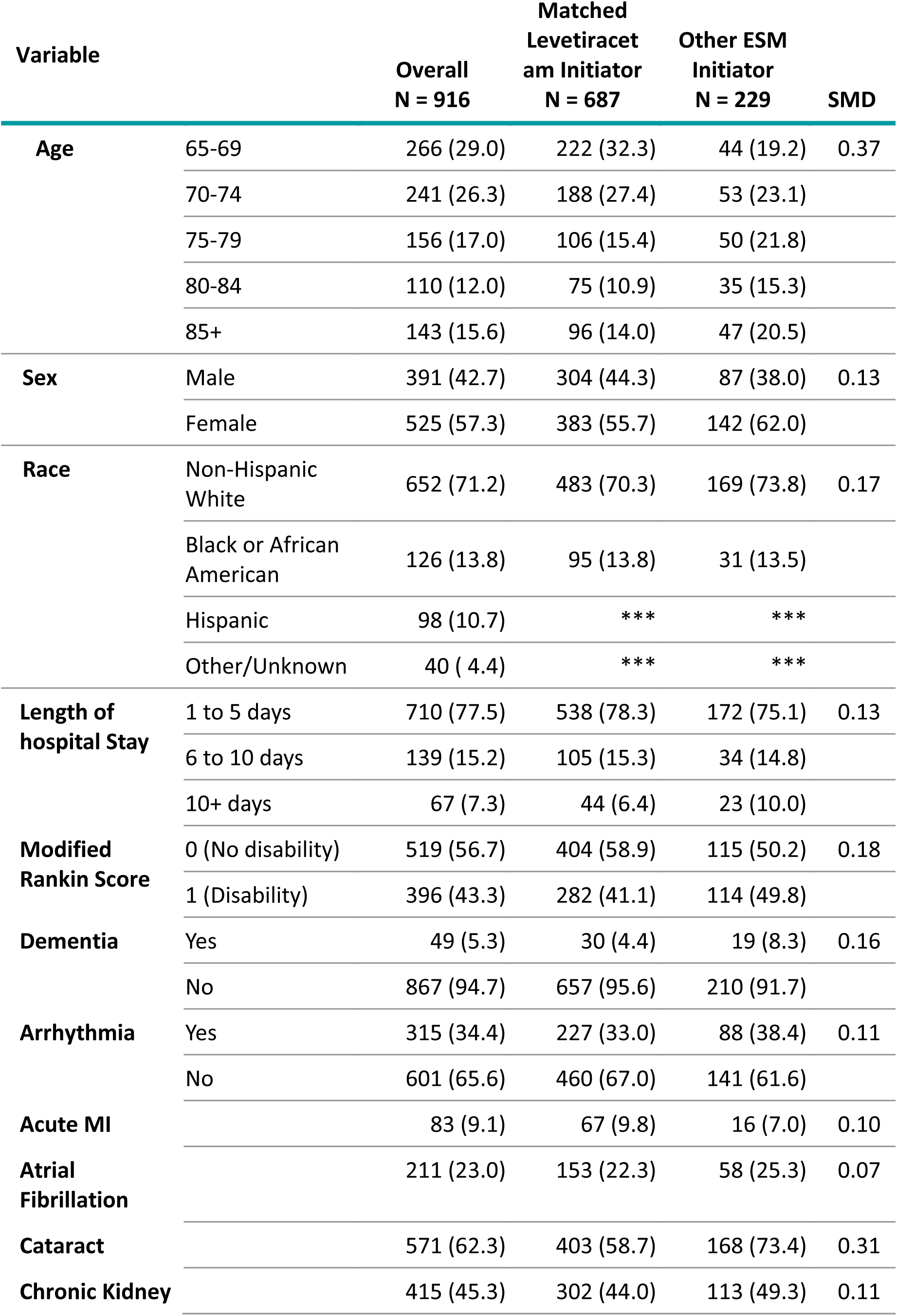
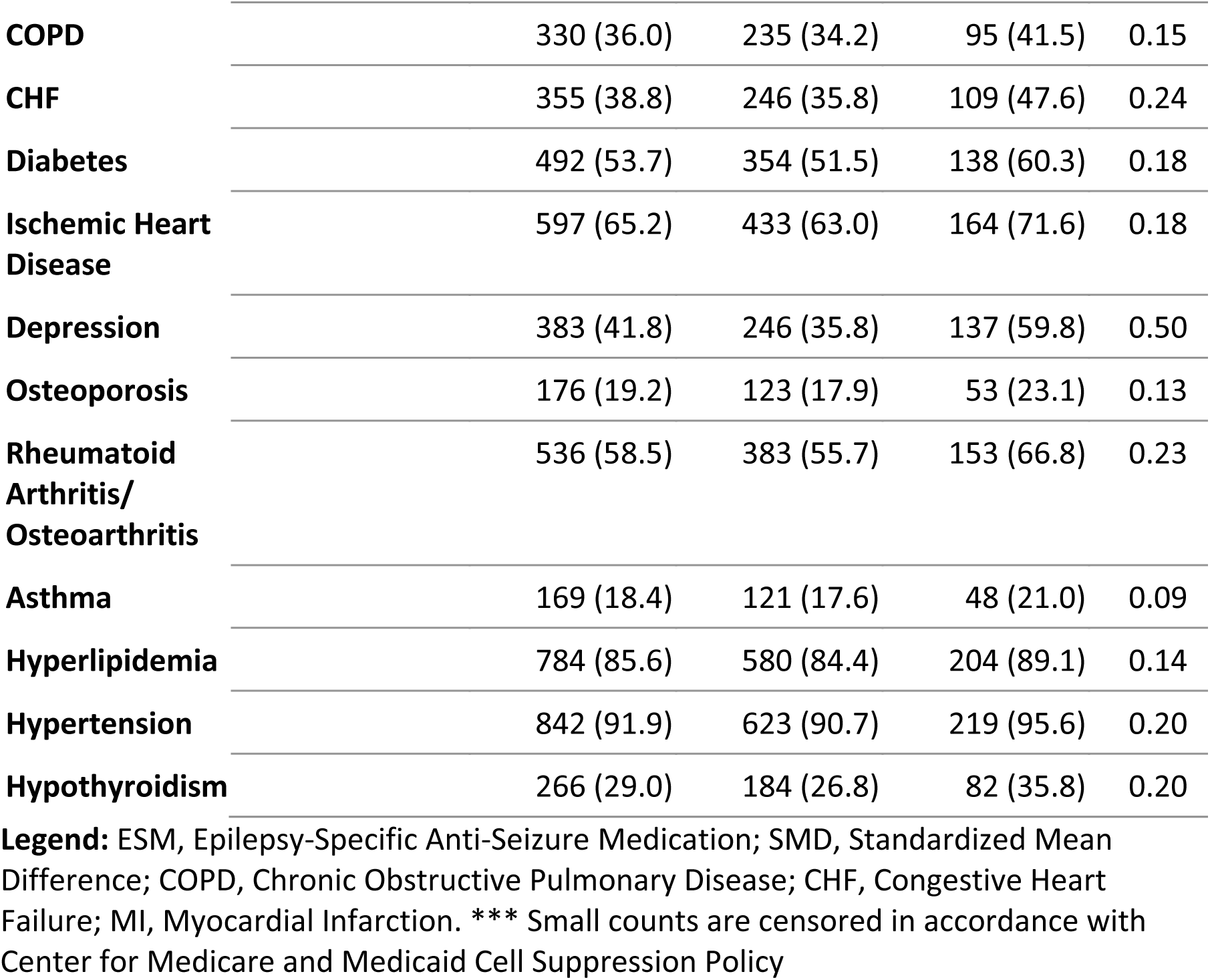
Sample Characteristics.

| Variable |  | Overall<br>N = 916 | Matched<br>Levetiracetam Initiator<br>N = 687 | Other ESM<br>Initiator<br>N = 229 | SMD |
| --- | --- | --- | --- | --- | --- |
| **Age** | 65-69 | 266 (29.0) | 222 (32.3) | 44 (19.2) | 0.37 |
|  | 70-74 | 241 (26.3) | 188 (27.4) | 53 (23.1) |  |
|  | 75-79 | 156 (17.0) | 106 (15.4) | 50 (21.8) |  |
|  | 80-84 | 110 (12.0) | 75 (10.9) | 35 (15.3) |  |
|  | 85+ | 143 (15.6) | 96 (14.0) | 47 (20.5) |  |
| **Sex** | Male | 391 (42.7) | 304 (44.3) | 87 (38.0) | 0.13 |
|  | Female | 525 (57.3) | 383 (55.7) | 142 (62.0) |  |
| **Race** | Non-Hispanic White | 652 (71.2) | 483 (70.3) | 169 (73.8) | 0.17 |
|  | Black or African American | 126 (13.8) | 95 (13.8) | 31 (13.5) |  |
|  | Hispanic | 98 (10.7) | *** | *** |  |
|  | Other/Unknown | 40 ( 4.4) | *** | *** |  |
| **Length of hospital Stay** | 1 to 5 days | 710 (77.5) | 538 (78.3) | 172 (75.1) | 0.13 |
|  | 6 to 10 days | 139 (15.2) | 105 (15.3) | 34 (14.8) |  |
|  | 10+ days | 67 (7.3) | 44 (6.4) | 23 (10.0) |  |
| **Modified Rankin Score** | 0 (No disability) | 519 (56.7) | 404 (58.9) | 115 (50.2) | 0.18 |
|  | 1 (Disability) | 396 (43.3) | 282 (41.1) | 114 (49.8) |  |
| **Dementia** | Yes | 49 (5.3) | 30 (4.4) | 19 (8.3) | 0.16 |
|  | No | 867 (94.7) | 657 (95.6) | 210 (91.7) |  |
| **Arrhythmia** | Yes | 315 (34.4) | 227 (33.0) | 88 (38.4) | 0.11 |
|  | No | 601 (65.6) | 460 (67.0) | 141 (61.6) |  |
| **Acute MI** |  | 83 (9.1) | 67 (9.8) | 16 (7.0) | 0.10 |
| **Atrial Fibrillation** |  | 211 (23.0) | 153 (22.3) | 58 (25.3) | 0.07 |
| **Cataract** |  | 571 (62.3) | 403 (58.7) | 168 (73.4) | 0.31 |
| **Chronic Kidney** |  | 415 (45.3) | 302 (44.0) | 113 (49.3) | 0.11 |

|  |  |  |  |  |
| --- | --- | --- | --- | --- |
| **COPD** | 330 (36.0) | 235 (34.2) | 95 (41.5) | 0.15 |
| **CHF** | 355 (38.8) | 246 (35.8) | 109 (47.6) | 0.24 |
| **Diabetes** | 492 (53.7) | 354 (51.5) | 138 (60.3) | 0.18 |
| **Ischemic Heart Disease** | 597 (65.2) | 433 (63.0) | 164 (71.6) | 0.18 |
| **Depression** | 383 (41.8) | 246 (35.8) | 137 (59.8) | 0.50 |
| **Osteoporosis** | 176 (19.2) | 123 (17.9) | 53 (23.1) | 0.13 |
| **Rheumatoid Arthritis/**<br>**Osteoarthritis** | 536 (58.5) | 383 (55.7) | 153 (66.8) | 0.23 |
| **Asthma** | 169 (18.4) | 121 (17.6) | 48 (21.0) | 0.09 |
| **Hyperlipidemia** | 784 (85.6) | 580 (84.4) | 204 (89.1) | 0.14 |
| **Hypertension** | 842 (91.9) | 623 (90.7) | 219 (95.6) | 0.20 |
| **Hypothyroidism** | 266 (29.0) | 184 (26.8) | 82 (35.8) | 0.20 |

**Legend:** ESM, Epilepsy-Specific Anti-Seizure Medication; SMD, Standardized Mean Difference; COPD, Chronic Obstructive Pulmonary Disease; CHF, Congestive Heart Failure; MI, Myocardial Infarction. \*\*\* Small counts are censored in accordance with Center for Medicare and Medicaid Cell Suppression Policy

Using a 0.2 threshold for defining a substantial difference based on standardized mean differences, age was found to be significantly associated with drug choice. Among individuals aged 65–69, 29% initiated Other ESMs compared with 32.3% of matched Levetiracetam initiators in the same age range.

### Association of ESM Choice and Outcomes

In the matched sample of 916 individuals, there were 298 seizure-like events, 465 ED visits, 382 hospital re-admissions and 137 mortality events within a follow-up period of 180 days. The average follow up time (to end of the follow-up window or death) was 163 days and the total follow up time was 410 person-years. Cumulative incidence curves for the outcomes are shown in Supplemental Materials – Figures 1-3. We report the results from the adjusted multivariable models investigating the association between ESM Choice and outcomes in Table 2-4. Survival curves for the outcomes are shown in Supplemental Materials – Figures 4-6. Among those who initiated ESMs other than Levetiracetam, the hazard of seizure-like events (given death has not occurred) was 37% lower than if they had initiated Levetiracetam, hazard ratio 0.63 [95% CI, 0.43, 0.91] (Table 2). The hazard of ED visits and readmission, given death had not occurred, was not different for the other ESM group compared with receiving LEV, with hazard ratios of 1.00 (95% CI: 0.80, 1.25) for ED visits and 0.98 (95% CI: 0.75, 1.28) for hospital readmission (Table 3 & 4).

**Figure 1.**
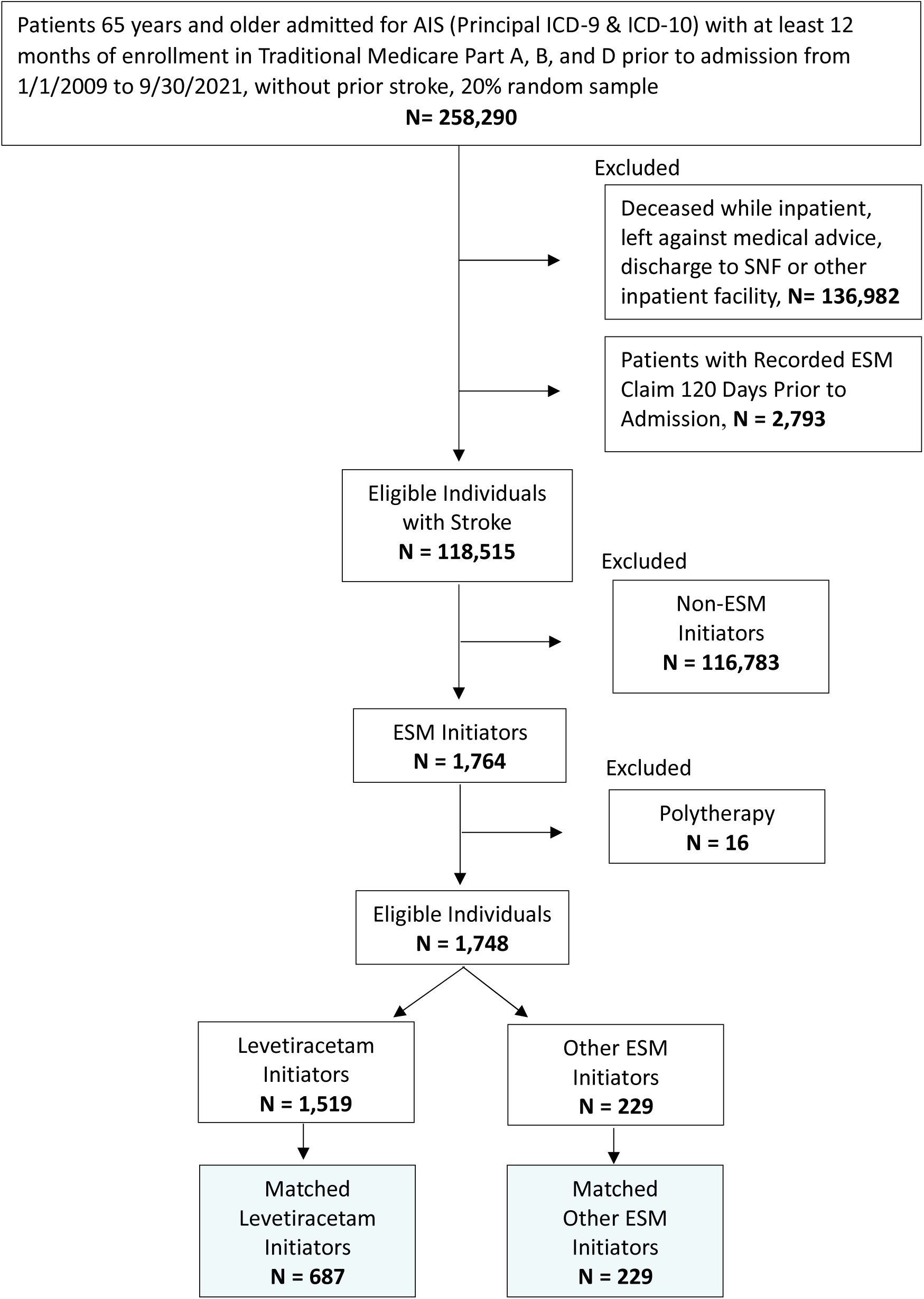
Sample Inclusion Criteria. AIS, Acute Ischemic Stroke; SNF Skilled Nursing Facility; ESM Epilepsy-Specific Anti-Seizure Medication

**Table 2.**
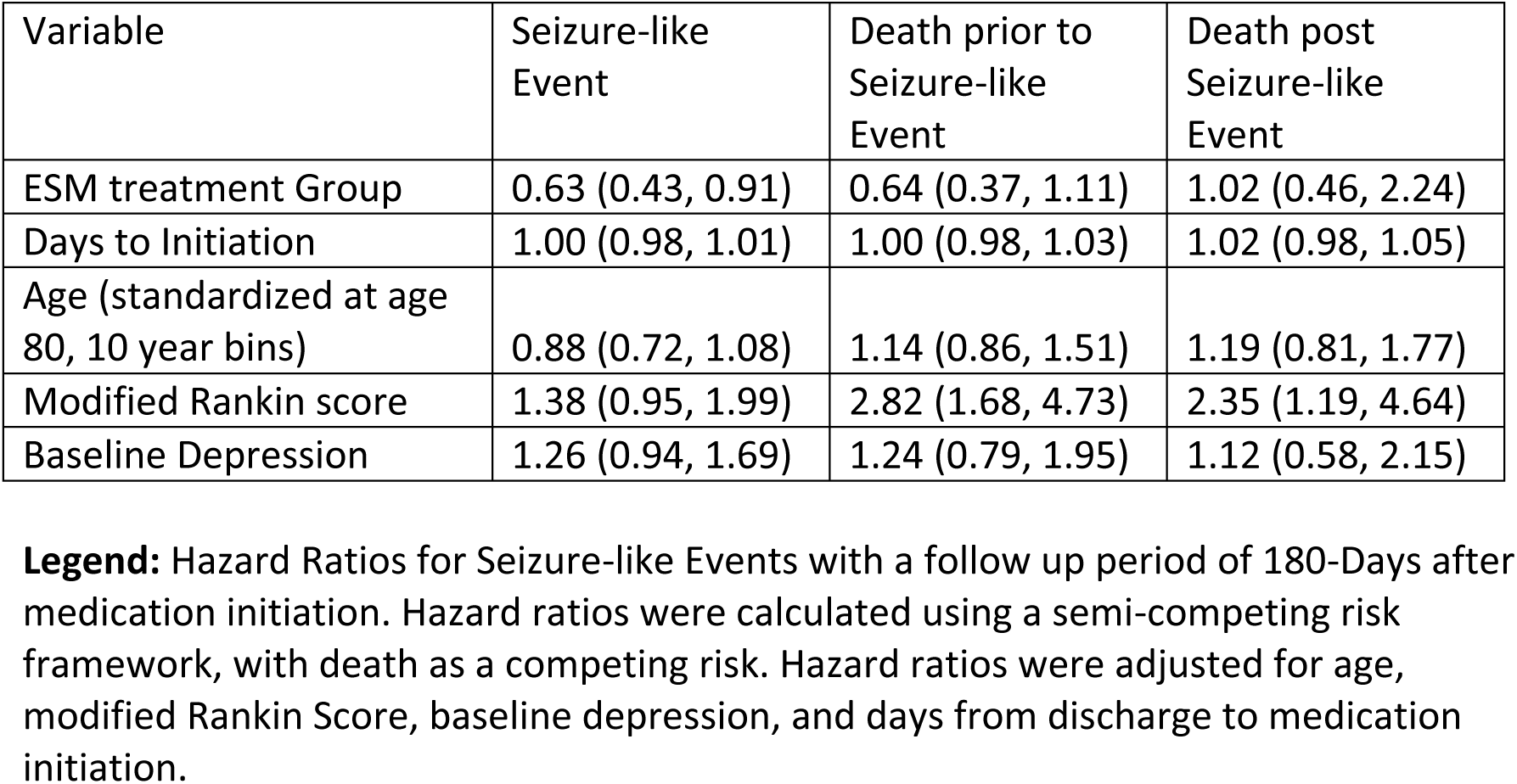
Hazard Ratios for Seizure-Like Events.

**Table 3.** Hazard Ratios for Emergency Department (ED) Visits.

| Variable | ED Visit | Death prior to ED Visit | Death post ED Visit |
| --- | --- | --- | --- |
| ESM treatment Group | 1.00 (0.80, 1.25) | 0.83 (0.44, 1.55) | 0.74 (0.43, 1.27) |
| Days to Initiation | 1.00 (0.99, 1.01) | 0.98 (0.95, 1.01) | 1.02 (1.00, 1.04) |
| Age (standardized at age 80, 10 year bins) | 1.02 (0.90, 1.17) | 1.34 (0.98, 1.85) | 1.04 (0.79, 1.38) |
| Modified Rankin score | 1.47 (1.16, 1.87) | 2.77 (1.51, 5.07) | 1.80 (1.11, 2.94) |
| Baseline Depression | 1.22 (0.99, 1.49) | 1.20 (0.70, 2.06) | 1.06 (0.67, 1.66) |

**Legend:** Hazard Ratios for ED Visits with a follow up period of 180-Days after medication initiation. Hazard ratios were calculated using a semi-competing risk framework, with death as a competing risk. Hazard ratios were adjusted for age, modified Rankin Score, baseline depression, and days from discharge to medication initiation.

**Table 4.**
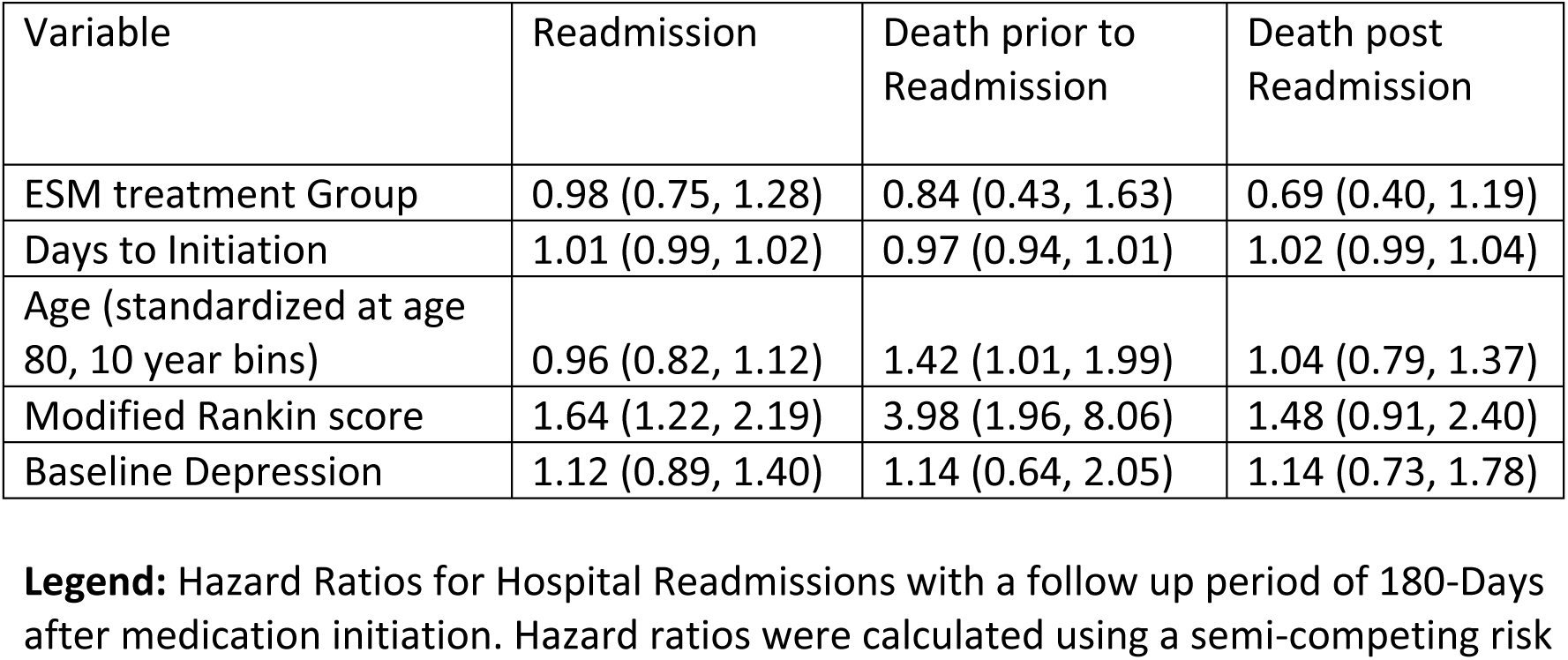
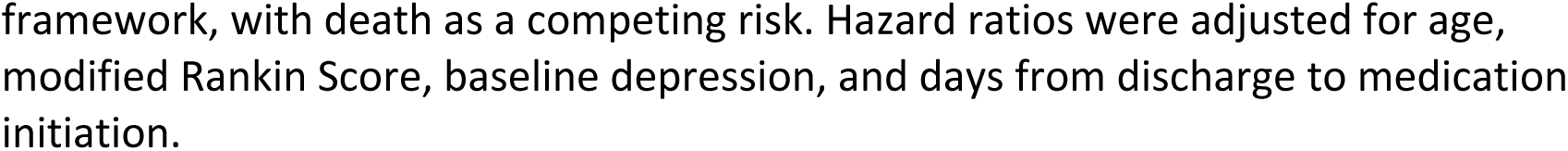
Hazard Ratios for Hospital Readmissions.

| Variable | Readmission | Death prior to Readmission | Death post Readmission |
| --- | --- | --- | --- |
| ESM treatment Group | 0.98 (0.75, 1.28) | 0.84 (0.43, 1.63) | 0.69 (0.40, 1.19) |
| Days to Initiation | 1.01 (0.99, 1.02) | 0.97 (0.94, 1.01) | 1.02 (0.99, 1.04) |
| Age (standardized at age 80, 10 year bins) | 0.96 (0.82, 1.12) | 1.42 (1.01, 1.99) | 1.04 (0.79, 1.37) |
| Modified Rankin score | 1.64 (1.22, 2.19) | 3.98 (1.96, 8.06) | 1.48 (0.91, 2.40) |
| Baseline Depression | 1.12 (0.89, 1.40) | 1.14 (0.64, 2.05) | 1.14 (0.73, 1.78) |

**Legend:** Hazard Ratios for Hospital Readmissions with a follow up period of 180-Days after medication initiation. Hazard ratios were calculated using a semi-competing risk
framework, with death as a competing risk. Hazard ratios were adjusted for age, modified Rankin Score, baseline depression, and days from discharge to medication initiation.

### Results of Secondary Pre-planned Analysis

In our analysis stratified by age, the hazard ratio for seizure-like events, given that death had not occurred was 0.54 (95% CI: 0.28, 1.04) for those less than or equal to 75 and 0.68 (95% CI: 0.45, 1.01) for individuals over 75 (Table 5). The results of the sensitivity analysis did not alter our conclusions (Supplemental Table S5 and S6).

**Table 5.**
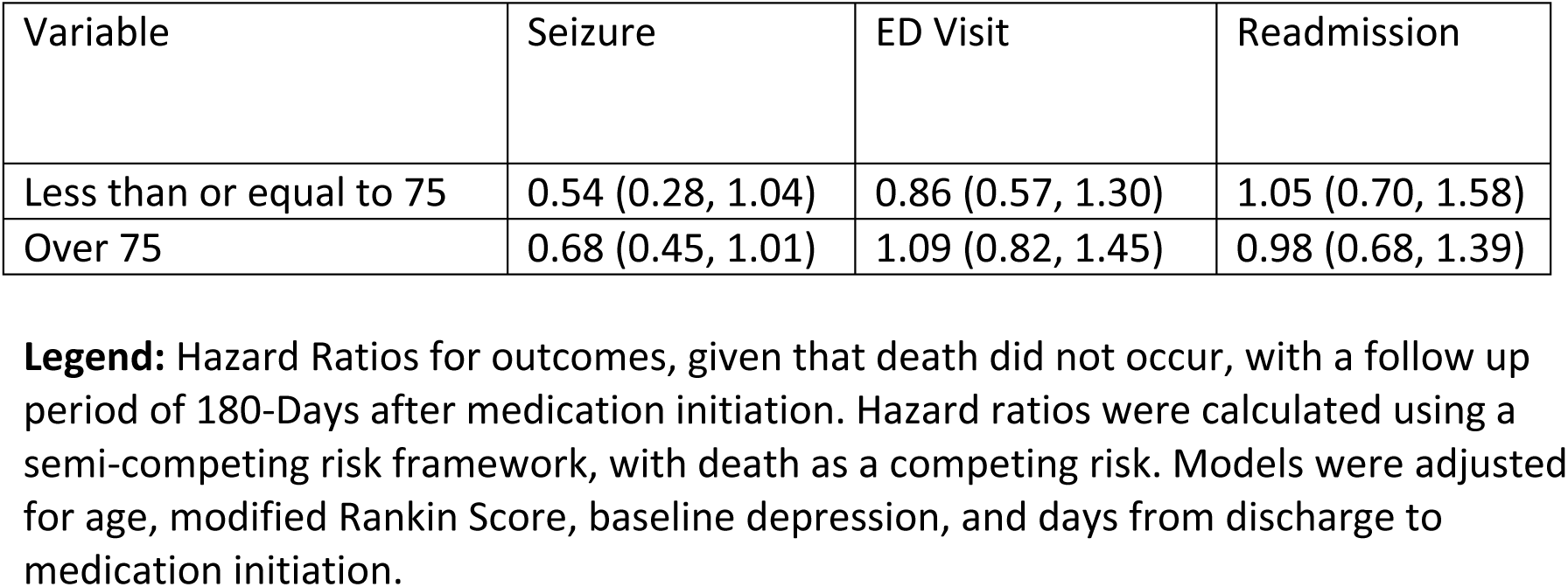
Hazard Ratios for Seizure-like Events, ED Visits, and Readmission, Stratified by Age Group.

## DISCUSSION

In a sample of Medicare beneficiaries hospitalized for acute ischemic stroke, we compared outcomes for matched individuals initiated on different ESMs. We found the risk of seizure-like events in those who initiated on Levetiracetam greater compared with other medications. In the context of insufficient guidance for seizure treatment in older patients, our study brings important information on post-stroke seizure control and ESM choice in this population.

We identified several previous randomized controlled studies on ESM choice for treatment of post-stroke seizures and epilepsy that are relevant to our study. Gilad et al. 2007 compared LTG (N = 32) with controlled-release carbamazepine (CR-CBZ) (N=32) and Consoli et a. 2012 compared LEV (n=52) with CR-CBZ (N = 54).^40,41^ A detailed review of these studies has been previously published by Brigo et al. 2018.^4^ In both previous studies of ESM choice, no difference was found in seizure freedom between groups, but both studies had a small number of patients included and thus may have been underpowered. In addition, randomized controlled trials are restrictive in their enrollment criteria and may have selection bias due to withdrawal from the trial. The average age of participants in Gilad et al. 2007 was 67.2 (SD 2.4) for LTG and 67.7 (SD 2.6) compared with our study population which had a median age of 74 (IQR 69 to 81) and participants ranging from 65 years old to over 85 years old. In Gilad et al. 2007, 3% of participants taking LTG and 31% of CR_CBZ withdrew from the trial due to adverse effects. Our study showed lower hazard of seizure-like events for other ESM initiators compared to receiving LEV.

We identified a population-based registry study on ESM choice in post-stroke epilepsy. Winter et al. 2022 followed 216 patients initiated on ESM monotherapy (LEV, LTG, LAC, VPA and eslicarbazepine (ESL)) for 12 months.^7^ The study reported lower seizure frequency in ESMs with selectivity to the slow-inactivated state of sodium channels (LAC, ESL) compared with other mechanisms of action.

Monotherapy with LEV has been previously found to have an effectiveness of more than 80% in preventing seizures.^42^ Unlike our study, other smaller studies did not find statistically significant differences in seizure-like events between post-stroke patients treated with LEV compared with other ESMs.^41^ These findings highlight the necessity of future guidelines for ESM therapy to distinguish older populations, considering the impact of increased seizure relapse in this group with an existing high risk of deficits and adverse outcomes.^43^ Moreover, previous randomized controlled studies present comparisons of ESM versus placebo, which does not help in drug type choice and can result in seizure relapse outcomes, as presented in our results.^44,45,46^

Stroke patients have a high risk of hospital readmission.^47–49^ A previous study found a total of more than 30% of stroke survivors being readmitted.^48^ The presence of a seizure diagnosis in this population has been reported as a contributing factor to both new hospitalizations and mortality risk.^47,49^ We did not identify any studies on ESM choice and differences in healthcare utilization (ED Visits and readmission). In our cohort, there was no difference in the ED visits, hospitalizations between patients treated with Levetiracetam compared to Other ESM.

Our study used a nationally representative sample to examine the relationship between ESM choice and outcomes, including more than 10 years of claims-based data. Currently available clinical data for ESM treatment choice for post-stroke epilepsy in the older population are not robust. Real-world evidence studies can help fill this knowledge gap and provide clinicians more information for ESM treatment options. Our sample permitted well measured outcomes for hospitalizations, ED visits and seizure outcomes, providing valuable, real-world information on ESM choice and outcomes.

### Limitations

The findings for our population sample might not be generalizable to groups not included in this study such as Medicare beneficiaries enrolled in Part C (Medicare Advantage), or patients not enrolled in Medicare prescription drug plans (Part D). Lastly, our sample of dementia patients initiated on other ESM was too small to complete stratification as initially planned.

This study excluded beneficiaries discharged to inpatient rehabilitation units or SNFs. Patients are discharged to these facilities usually present with variable disabilities and would benefit from assessment in future studies.^50^ We also focused on subacute outcomes of ESM treatment on acute ischemic stroke patients. Later studies could explore chronic outcomes, especially when considering the different patterns of ESM use and adverse effects among this population.^9^

Due to existing limitations in the dataset, some residual confounding factors could not be controlled. We were not able to adjust for factors associated with stroke severity and seizure outcomes, such as stroke territory. We found that as covariates were added incrementally to the model specification, the differences between treatment groups became larger, as indicated by lower hazard ratios (Table S6). Additional variables, could reduce variance and improve precision, but would not necessarily enhance the validity of the model because of their strong correlation with variables already included the model. After the adjustments made, residual confounding from unmeasured factors is likely minimal.

While the use of claims-based data is reliable, it also has limitations such as missing data and a lack of granular seizure outcome data. Data are also absent on seizure frequency, seizure type, date of last seizure episode, or indication for ESM treatment. We tried to address this last point by selecting ESMs to include in this study that are primarily indicated for treatment of seizures. Lastly, certain ESM presented with too small groups and so we did not have the power to see small differences between groups.

## Data Availability

The data supporting this study's findings were collected by The Centers for Medicare & Medicaid Services (CMS) and were made available by CMS with no direct identifiers. All results were aggregated following CMS Cell Suppression Policies. Restrictions apply to the availability of these data, which were used under license for this study. Medicare data are available through CMS with their permission.

## Disclosures

J.D.B. completed the data analysis, drafted and edited the. R.C.M. drafted, edited, and revised the manuscript. S.S. and M.S. were involved with study design and statistical analysis. B.W. and L.S. were involved in the study conceptualization and revised the manuscript. J.P.N. facilitated data access and revised the manuscript. S.H. was involved in study design, supervising the study development, and revising the manuscript.

L.M.V.R.M. was involved in study design and conceptualization, obtaining data access, and supervising the study development and revising the manuscript.

